# Effects of THAM Nasal Alkalinization on Airway Microbial Communities: A Pilot Study

**DOI:** 10.1101/2021.02.14.21251657

**Authors:** Zachary M. Holliday, Janice L. Launspach, Lakshmi Durairaj, Pradeep K. Singh, Joseph Zabner, David A. Stoltz

## Abstract

**Objectives:** In cystic fibrosis (CF), loss of CFTR-mediated bicarbonate secretion reduces the airway surface liquid (ASL) pH causing airway host defense defects. Aerosolized sodium bicarbonate can reverse these defects, but its effects are short-lived. Aerosolized tromethamine (THAM) also raises the ASL pH but its effects are much longer lasting. In this pilot study, we tested the hypothesis that nasally administered THAM would alter the nasal bacterial composition in adults with and without CF.

**Methods:** Subjects (n=32 total) received intranasally administered normal saline or THAM followed by a wash out period prior to receiving the other treatment. Nasal bacterial cultures were obtained prior to and after each treatment period.

**Results:** At baseline, nasal swab bacterial counts were similar between non-CF and CF subjects, but CF subjects had reduced microbial diversity. Both nasal saline and THAM were well-tolerated. In non-CF subjects, nasal airway alkalinization decreased both the total bacterial density and the gram-positive bacterial species recovered. In both non-CF and CF subjects, THAM decreased the amount of *C. accolens* detected, but increased the amount of *C. pseudodiphtheriticum* recovered on nasal swabs. A reduction in *S. aureus* nasal colonization was also found in subjects who grew *C. pseudodiphtheriticum*.

**Conclusions:** This study shows that aerosolized THAM is safe and well-tolerated and that nasal airway alkalinization alters the composition of mucosal bacterial communities.

## INTRODUCTION

In the CF airways, loss of Cl^-^ and HCO_3_^-^ secretion, by the cystic transmembrane conductance regulator (CFTR), causes multiple host defense defects against inhaled and/or aspirated bacteria (1). This leads to recurrent and chronic airway infections and inflammation. While antibiotic use and CFTR modulator therapy have markedly improved outcomes in CF, CF remains a life-shortening disease and available data suggest that chronic airway infections persist even after treatment with highly effective CFTR modulator therapy (1-5).

The airways are lined with a thin layer of liquid termed the airway surface liquid (ASL), whose composition and volume are tightly regulated (6). The ASL is the first line of defense for the respiratory tract and contains antimicrobial factors, which are important for bacterial killing, and mucus, which plays an important role in mucociliary transport (MCT) (6, 7). Newborn CF pigs manifest at least two host defense defects, including impaired antimicrobial factor-mediated bacterial killing and defective mucociliary transport (8-14). Both defects are related to loss of CFTR-mediated HCO_3_^-^ secretion which leads to a more acidic ASL pH in newborn CF pigs, CF rats, and infants with CF (15-17). In primary cultures of pig and human CF airway epithelia, similar findings are present (18-21). However, with disease progression the effects of CFTR loss on ASL pH is less clear (16, 22, 23).

One approach to restoring airway host defenses in CF would be to raise the ASL pH. In the newborn CF pig airway, airway alkalinization with aerosolized sodium bicarbonate (NaHCO_3_) restored bacterial killing, suggesting that airway alkalinization might be a therapeutic approach in CF (15). However, this response was transient and ASL pH returned to normal levels within 30 minutes (24). Aerosolized tromethamine (THAM or tris[hydroxymethyl]aminomethane acetate), another alkalinizing agent, also raised the ASL pH but its effects were much longer lasting than NaHCO_3_ and persisted for at least 2 hours (24). THAM is an FDA-approved buffer which has been used intravenously for metabolic acidosis in the critical care setting and as an excipient in a number of medications, including inhaled prostacyclin and nasal ketorolac (25, 26). Therefore, aerosolized THAM could represent a novel therapeutic for CF-related airway disease.

While we know that THAM raises ASL pH, we do not know whether ASL alkalinization will alter bacterial density, how much of bacterial killing *in vivo* is pH-dependent, whether raising pH might have negative consequences, or how communities of bacteria will respond to alkalinization. This pilot study is the next key step and we tested the hypothesis that nasally administered THAM would alter the nasal bacterial composition in adult subjects, with and without CF. We chose to focus on the upper airways (nose) because aerosolized delivery to this region is straightforward, has minimal risks, microbiological sampling of the nasal airways is relatively easy, and the biology of the upper and lower airways is similar (27, 28).

## RESULTS

### Baseline subject characteristics

A total of 32 subjects were enrolled in the study (16 non-CF and 16 CF subjects) (**Table 1**). Age, sex, and race were similar between the two groups. Eighty-one percent of CF subjects had at least one *ΔF508-CFTR* allele present. Prior to study enrollment, the average FEV1 % predicted was 73% in CF participants. No subjects were lost to follow-up. Treatment compliance was excellent with non-CF and CF subjects completing 97% and 99% of aerosolized treatments, respectively. Both aerosolized saline and THAM were very well-tolerated. Adverse events were limited with 4 events noted during saline treatment (1 cough, 2 shortness of breath, 1 chest tightness) and 2 during THAM treatment (1 sneezing, 1 back pain) (**Supplemental Table 1**).

**Table 1.**

| Subject Demographics |  |  |
| --- | --- | --- |
|  | non-CF | CF |
| Number of subjects | 16 | 16 |
| Age (yrs) | 33 | 29 |
| Male/female | 5/11 | 9/7 |
| Caucasian/white | 15 (94%) | 16 (100%) |
| ΔF508 heterozygotes | N/A | 8 (50%) |
| ΔF508 homozygotes | N/A | 5 (31%) |
| FEV<sub>1</sub> (% predicted) | N/A | 73% |

In preliminary studies, we found that both total amount and types of bacteria recovered from the nares were stable over time (**Supplemental Figure 1A-D**). Prior to any treatments, nasal mucosal swabs recovered similar numbers of bacteria from non-CF and CF subjects (4.03 ± 0.25 vs. 3.91 ± 0.24 log_10_ CFU/ml, P = 0.731) (**Supplemental Figure 2A**). In the current study, bacterial diversity, represented by the number of different bacterial species isolated, was reduced in CF compared to non-CF subjects (mean number of different species present: 2.6 ± 0.3 vs. 3.9 ± 0.5, P = 0.023) (**Supplemental Figure 2B**). In both non-CF and CF subjects, the majority of bacteria isolated were gram-positive species (**Supplemental Table 2**). Gram-negative bacterial species were not frequently isolated from either non-CF or CF subjects, but were more commonly cultured from CF nasal mucosa with the most common in CF being *P. aeruginosa, Achromobacter xylosoxidans*, and *Serratia marcescens. P. aeruginosa* was only cultured from the nares of CF subjects.

**Table 2.**
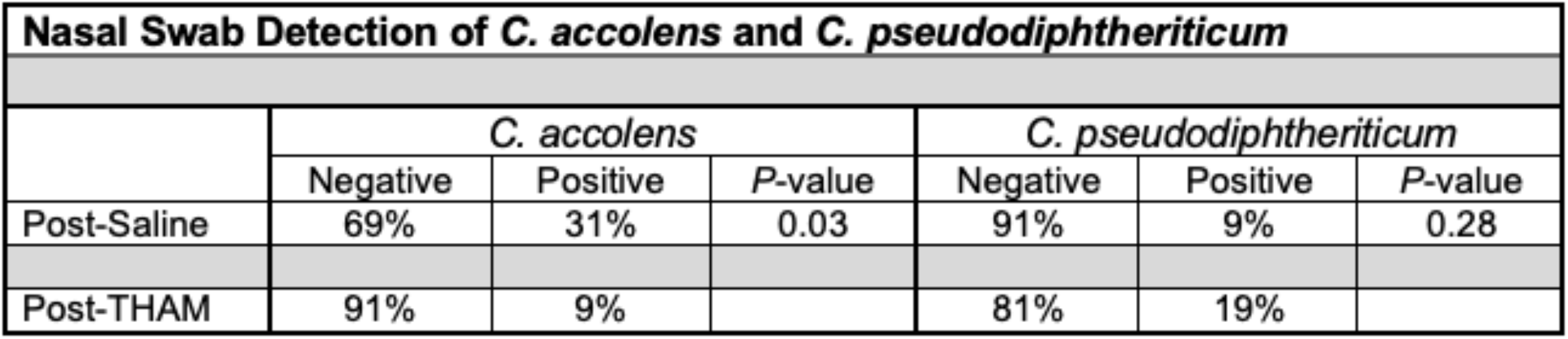

**Figure 1.**
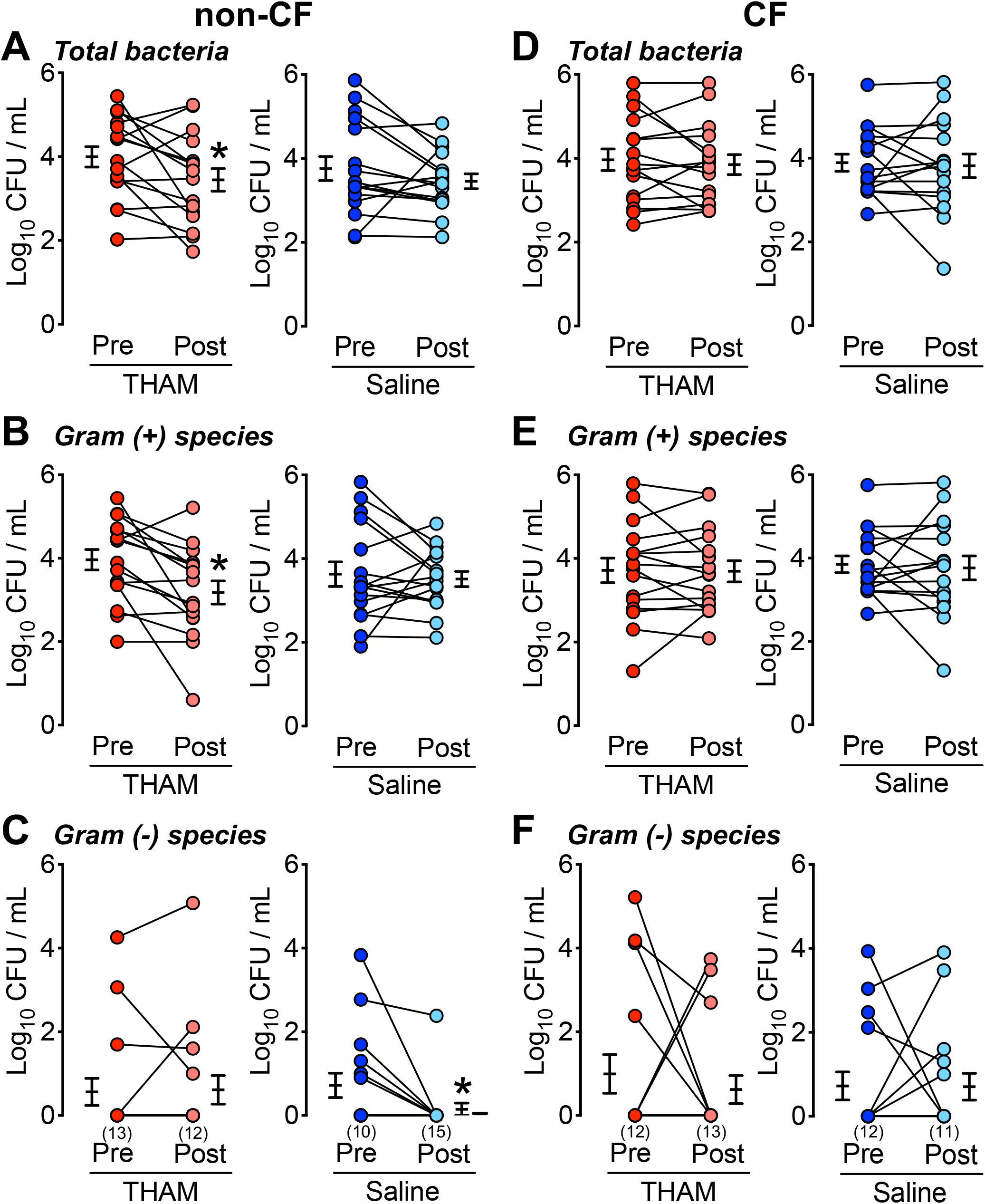
The Effects of Airway Alkalinization on Nasal Bacteria in Non-CF and CF Subjects. (**A**) Total bacteria, (**B**) Gram-positive bacteria, and (**C**) Gram-negative bacteria cultured from nasal swabs before and after nasal aerosolization with THAM (red symbols) or saline (blue symbols) in non-CF subjects. (**D**) Total bacteria, (**E**) Gram-positive bacteria, and (**F**) Gram-negative bacteria cultured from nasal swabs before and after nasal aerosolization with THAM (red symbols) or saline (blue symbols) in CF subjects. Each data point indicates nasal swab data from a different subject. Connected symbols represent individual subjects before and after treatment. n = 16 subjects for non-CF and 16 subjects for CF. Numbers in parenthenses represent the number of subjects with a bacterial count of “0” and have superimposed data points. Horizontal bars represent mean ± SEM. * denotes P < 0.05.

**Figure 2.**
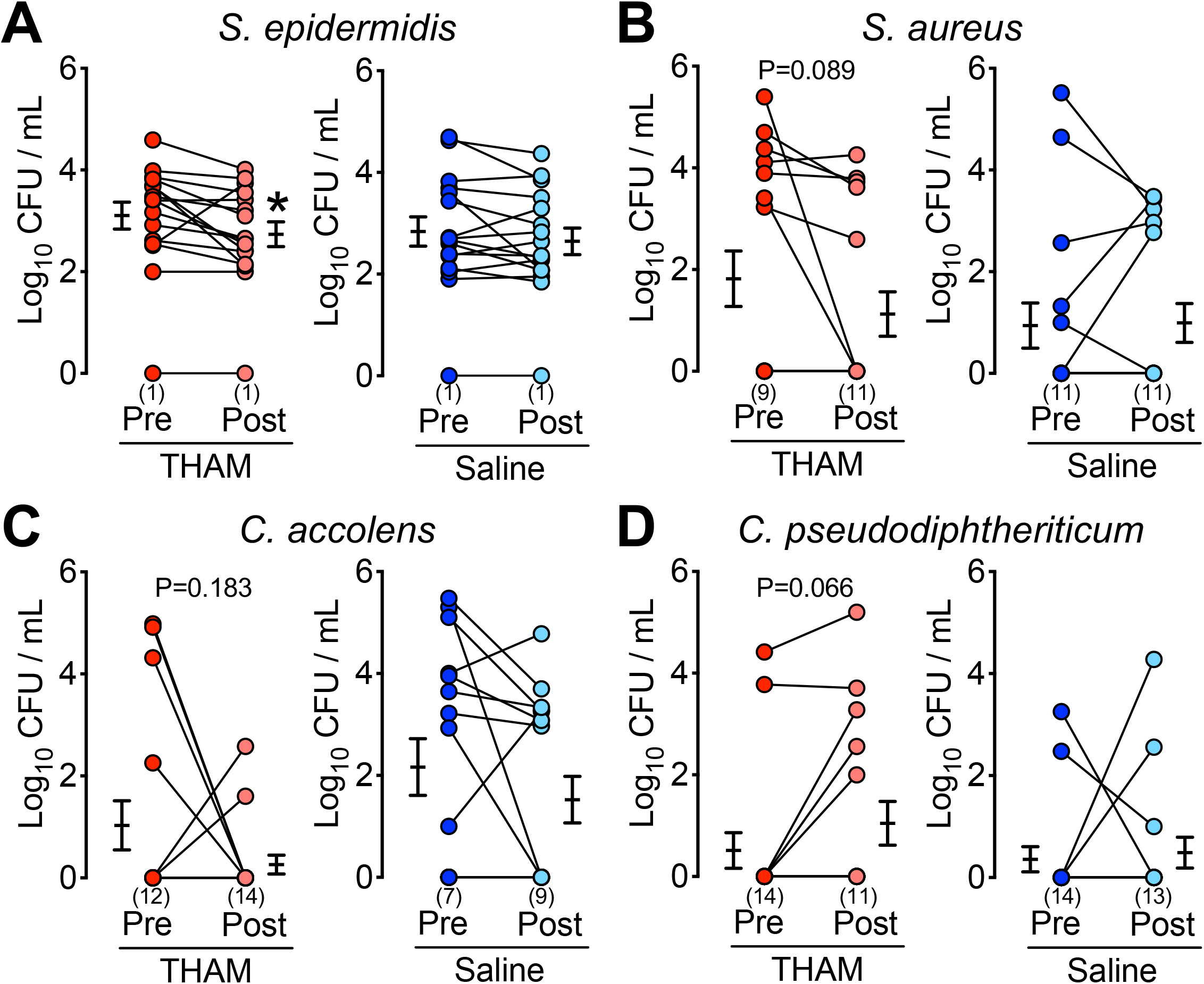
The Effects of Airway Alkalinization on the Most Common Gram-Positive Nasal Bacterial Species in Non-CF Subjects. Nasal bacterial counts for (**A**) *S. epidermidis*, (**B**) *S. aureus*, (**C**) *C*. accolens, and (**D**) *C. pseudodiphtheriticum*. Bacteria recovered before and after nasal aerosolization with THAM (red symbols) or saline (blue symbols) in non-CF subjects. Each data point indicates nasal swab data from a different subject. Connected symbols represent individual subjects before and after treatment. n = 16 subjects. Numbers in parenthenses represent the number of subjects with a bacterial count of “0” and have superimposed data points. Horizontal bars represent mean ± SEM. * denotes P < 0.05.

### Nasal airway alkalinization decreases gram-positive bacterial species in non-CF subjects

Aerosolized THAM reduced the total number of bacteria recovered from the nares of non-CF subjects (**Figure 1A**), and this effect was primarily due to a decrease in gram-positive bacteria (3.96 ± 0.98 vs 3.18 ± 1.11 log_10_ CFU/ml, P = 0.005) (**Figures 1B and 1C**). Nasal saline aerosolization had no effect on the counts of total and gram-positive bacteria recovered from the nares of non-CF subjects (**Figure 1A and 1B**). Unexpectedly, nasal saline decreased the total number of gram-negative bacteria (0.86 ± 1.20 vs. 0.15 ± 0.60 log_10_ CFU/ml, P = 0.020) (**Figure 1C**), although most participants had no gram-negative or relatively low levels of these organisms.

In CF subjects, aerosolized THAM or saline had no effect on total bacterial counts or the total number of gram-positive or gram-negative bacteria recovered (**Figures 1D-F**). Prior to THAM treatment, *P. aeruginosa* was recovered from the nares of 4 people with CF. Following THAM treatment, *P. aeruginosa* was only recovered from 1 of these 4 subjects and in this remaining subject THAM decreased the amount of *P. aeruginosa* (4.18 vs. 2.70 log_10_ CFU/ml). Saline had no significant effect on *P. aeruginosa* recovery.

In non-CF subjects, the four most common gram-positive organisms detected on nasal swabs at baseline were *S. epidermidis, S. aureus, C. accolens*, and *C. pseudodiphtheriticum*. Aerosolized THAM, but not saline, treatment decreased the amount of *S. epidermidis* (3.11 ± 1.06 vs. 2.74 ± 0.98 log_10_ CFU/ml, P = 0.031), *S. aureus* (1.82 ± 2.18 vs. 1.12 ± 1.75 log_10_ CFU, P = 0.089), and *C. accolens* (1.03 ± 1.93 vs. 0.26 ± 0.74 log_10_ CFU, P = 0.183) cultured from nasal swabs (**Figures 2A-C**). In contrast, we cultured higher amounts of *C. pseudodiphtheriticum* following THAM treatment (0.51 ± 1.40 vs. 1.05 ± 1.72 log_10_ CFU/ml, P = 0.066 (**Figure 2D**).

Since the number of subjects in each group with *C. accolens* or *C. pseudodiphtheriticum* was relatively low, we combined the values from non-CF and CF subjects and reanalyzed the data. A similar pattern was observed with THAM treatment decreasing the amount of *C. accolens*, but increasing the amount of *C. pseudodiphtheriticum* cultured from nasal swabs (**Figures 3A and 3B**). We also found that following THAM treatment, *C. accolens* was less frequently detected on nasal swabs (9% vs. 31% for saline treatment, P = 0.03) while there was a trend towards increased detection of *C. pseudodiphtheriticum* (9% vs. 19% for THAM treatment, P = 0.28) (**Table 2**). Finally, the presence of *C. pseudodiphtheriticum*, but not *C. accolens*, in both non-CF and CF subjects, was associated with a significantly lower rate of *S. aureus* detection on nasal swab cultures (**Figure 3C**).

**Figure 3.**
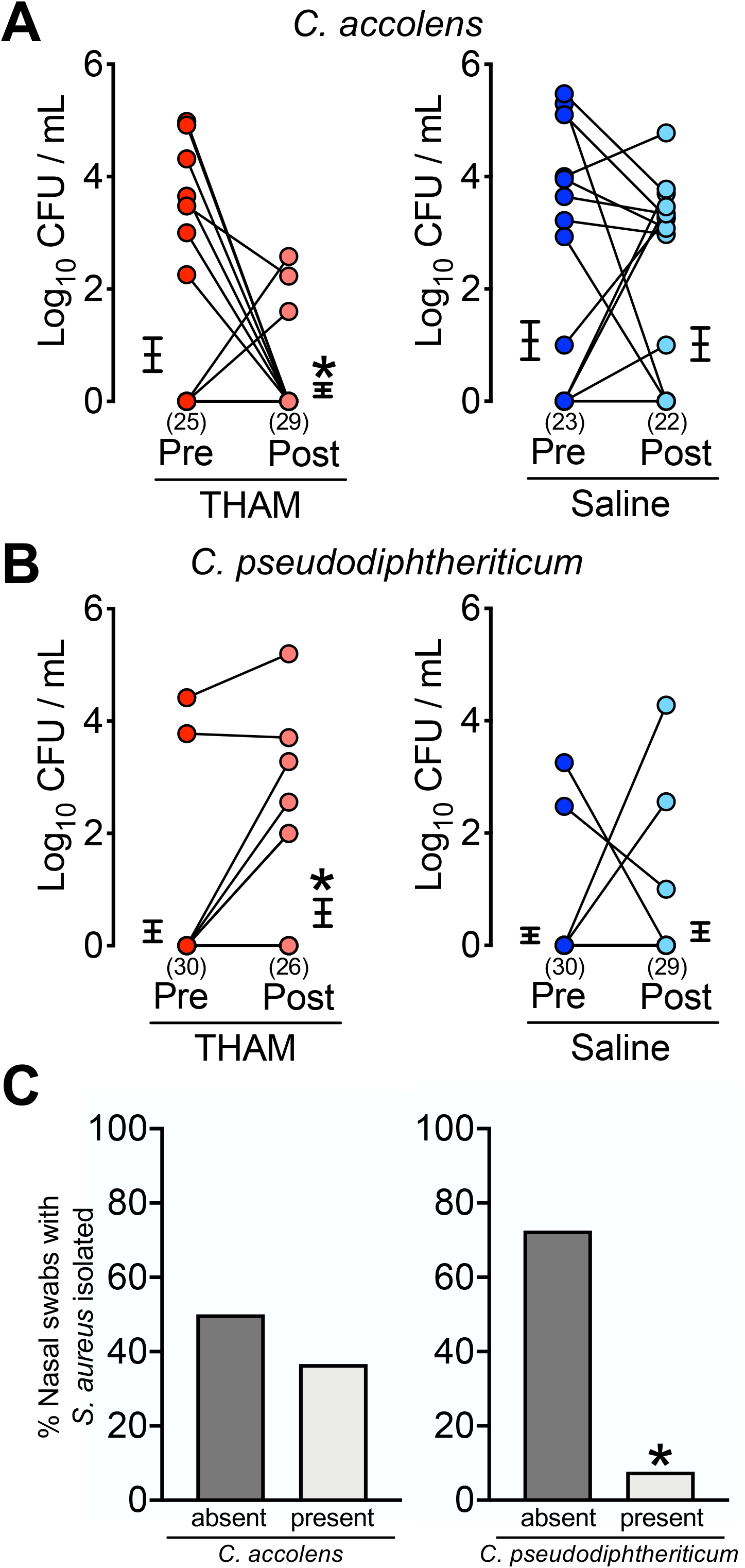
THAM’s Effects on *C. accolens, C. pseudodiphtheriticum*, and *S. aureus* in non-CF and CF Subjects. Nasal bacterial counts for (**A**) *C. accolens* and (**B**) *C. pseudodiphtheriticum*, in non-CF and CF subjects. Bacteria recovered before and after nasal aerosolization with THAM (red symbols) or saline (blue symbols) in non-CF and CF subjects. Each data point indicates nasal swab data from a different subject. Connected symbols represent individual subjects before and after treatment. n = 32 subjects. Numbers in parenthenses represent the number of subjects with a bacterial count of “0” and have superimposed data points. Horizontal bars represent mean ± SEM. * denotes P < 0.05. (**C**) Percentage of nasal bacterial swab samples positive for *S. aureus* in the absence and presence of *C. accolens*- or *C. pseudodiphtheriticum* positive samples. * denotes P < 0.05.

## DISCUSSION

In this study investigating the effects of nasal alkalinization, we found that THAM was well-tolerated and altered the types of bacteria cultured from the nasal mucosa of both non-CF and CF subjects. We had four main conclusions. First, at baseline, non-CF and CF subjects had similar total numbers of bacteria detected on nasal swabs, but there was less microbial diversity in CF. Second, airway alkalinization with THAM preferentially reduced the total number of gram-positive bacteria recovered in non-CF subjects, but had less of an effect in CF. Third, following airway alkalinization, nasal swab cultures were less frequently positive for *C. accolens*. Finally, the presence *C. pseudodiphtheriticum* was associated with much less frequent recovery of *S. aureus*.

We found that THAM’s effects were most significant for gram-positive species. This is likely because, in part, gram-negative species are uncommonly present in the upper airways, with less than 10% of healthy individuals nasally colonized with gram-negative bacteria (31). We also observed low rates of gram-negative bacteria in the nares of non-CF subjects. Although this was only a small pilot study, we did observe that THAM treatment decreased *P. aeruginosa* detection from the nares of CF subjects. Whether this effect is reproducible in a larger patient population or in the lower airways is not known, but would be an important endpoint for future studies of airway alkalinization of individuals with CF colonized with *P. aeruginosa*.

An interesting finding was that airway alkalinization altered the types and amounts of bacteria recovered from the nares of non-CF and CF subjects. Specifically, following THAM treatment, decreased amounts of *C. accolens* were cultured, while increased amounts of *C. pseudodiphtheriticum* were present on nasal swab cultures. While we did not investigate the underlying mechanism several potential explanations exist. First, THAM might directly affect the growth of certain bacterial species. THAM could inhibit the growth of *C. accolens*, but promote the growth of *C. pseudodiphtheriticum*. Second, it is possible that airway alkalinization alters the nasal mucosa’s innate defenses. The bacterial killing activity of many antimicrobial factors present on the airway surface is affected by pH (32). Airway alkalinization could enhance or diminish antimicrobial factor killing of different bacterial species and alter the bacterial community composition. Finally, alkalinization and environmental changes might select for certain bacterial species which then shift the remaining microbial community. *C. accolens* has been shown to have a symbiotic effect when cultured with *S. aureus*, while *C. pseudodiptheriticum* negatively affects the growth of *S. aureus* (33, 34). Interestingly, an earlier study found that nasal inoculation with *C. pseudodiphtheriticum* significantly decreased *S. aureus* nasal carriage (35). Thus, THAM might be selecting for *C. pseudodiphtheriticum* which then reduces the amount of *Staphylococcus* and *C. accolens*. This might represent an example of how changing the local host enviroment favors the colonization of different bacterial species.

Although we did not observe that nasal THAM alkalinization decreased the frequency of *S. aureus* detection, there was a reduction in the amount of *S. aureus* cultured from nasal swabs. It is possible that with a more prolonged duration of akalinization, THAM might represent a novel therapeutic strategy for *S. aureus* decolonization. Nasal THAM administration might be a simple, cost-effective, and efficacious approach for nasal *S. aureus* decolonization, but will require future studies designed to test its effectiveness. The mechanism underlying THAM’s effects are likely due to increased activity of antimicrobial-factors. Since resistance to multiple innate factors is difficult to develop, this approach would be less likely to cause bacterial resistance.

This study has both advantages and limitations. Advantages include: a) We studied both non-CF and CF human subjects. b) A cross-over study design was utilized allowing for comparisons to be made on the same subject. c) An FDA-approved formulation of THAM was used. Limitations include: a) This was a pilot study, at a single-center, with a limited number of subjects. b) We only studied the upper (nasal) airways. However, this allowed us to easily administer treatments and sample the airways. c) The study duration was short. We do not know the long-term effects of airway alkalinization on airway bacterial colonization. d) Bacterial recovery with nasal swabs can be variable, but we standardized the approach based upon currently accepted techniques. e) We did not measure the effects of THAM on nasal pH. However, in an earlier study we found that the effect lasted close to 6 hours (24). Thus, we likely had prolonged airway alkalinization by administering the treatment three times per day.

In conclusion, nasal THAM administration was safe and well-tolerated in both non-CF and CF subjects. THAM treatment decreased the number of gram-positive bacteria cultured from the nasal mucosa of non-CF subjects, but had less of an effect in CF. In both non-CF and CF subjects, alkalinization was associated with reduced *C. accolens* detection and a trend towards greater rates of *C. pseudodiphtheriticum* nasal carriage. Future studies will be required to further understand the mechnaisms underlying the changes in the bacterial communities. Finally, airway alkalinization with THAM might represent a potential therapeutic approach in early CF.

## METHODS

### Subjects

All subjects were recruited and enrolled at the University of Iowa Hospital. We enrolled people with and without CF age 16 or greater, who were able to provide written consent. A CF diagnosis required one or more features consistent with CF and one or more of the following criteria: a) sweat chloride ≥ 60 mEq/L; b) two mutations in *CFTR*; or c) an abnormal nasal potential difference (29). Inclusion criteria for CF subjects included resting O_2_ saturation > 90% on room air and clinically stable for at least 14 days prior to Day 1 of the study. CF subjects were recruited from the CF clinic. Exclusion criteria were pregnancy, tobacco use, recreational drug use, use of any investigational study drug within 28 days, or clinical findings consistent with a CF pulmonary exacerbation or flare up of seasonal allergic rhinitis. Baseline pulmonary function testing (PFT) was available for only the CF subjects.

### Study approval

The University of Iowa IRB as well as the FDA approved the studies. A written informed consent was received from participants prior to inclusion in the study. The clinical trial registration number for this study is NCT03078088.

### Study Design and Protocol

This pilot study was a prospective, randomized, cross-over design. Subjects were randomized to receive either normal saline (0.9%, Baxter Health Care, Mundelein, IL) (250 μl/dose) or THAM (3.6%, pH 8.6, Pfizer) (250 μl/dose) 3 times a day for 4 days. After a 10 – 14 day wash out period, subjects then received the other treatment for 4 days. An Accuspray syringe (Becton Dickinson, Franklin Lakes, NJ) was used to aerosolize the solutions onto the nasal epithelium, as previously described (30). Subjects received prefilled syringes from Nucara Pharmacy (Waterloo, IA). These treatments were self-administered by the study subjects. Bacterial cultures of the anterior nares were obtained before and after each round of treatment.

### Nasal Swab Sampling and Microbiology

Nasal bacteria were recovered by gently inserting a flexible minitip flocked swab (BD ESwab, cat# 220532) into the right nare 6 cm and completely rotating the swab three times. The swab was carefully removed, placed in 1 ml of media, and vortexed for 20 seconds. The same procedure was performed in the left nare with a different swab. Using standard microbiological techniques, the media was plated for bacterial quantification and identification via MALDI-TOF mass spectrometry. Data from the right and left nares were combined for each subject at the different time points.

### Statistical Analyses

Data are presented as points from individual donors with mean ± SEM indicated by bars. For statistical analysis, we used a two sample t-test, Mann-Whitney test, Wilcoxon signed-rank test, or a 1-way ANOVA. Bacterial counts were log-transformed. Prior to log transformation, a value of 1 was added to all CFU data. Differences were considered statistically significant at P < 0.05.

## Data Availability

The data are available from Drs. Zabner and Stoltz.

## TRANSPARENCY DECLARATION

### Conflict of Interest

Drs. Zabner and Stoltz have a patent application pending related to this work.

### Funding

This research was supported, in part, by the NIH (HL136813, HL091842, and HL007638), and the Cystic Fibrosis Foundation (Iowa Cystic Fibrosis Foundation Research Development Program). This study was also supported by the National Center For Advancing Translational Sciences of the NIH under award number UL1TR002537.

## Acknowledgements

The authors thank the people with CF for their participation. We thank Linda Boyken, Nick Gansemer, Mal Stroik, and Anne Vincent for excellent assistance. We thank the members of the data and safety monitoring board, Drs. Doug Hornick and Anthony Fischer, for their valuable service to this study.

## Access to Data

The data are available from Drs. Zabner and Stoltz.

## Contributions

ZMH, LD, PKS, JZ, DAS conception and design of research; ZMH, JLL performed experiments and data acquisition; ZMH, JZ, DAS analyzed data; ZMH, LD, PKS, JZ, DAS interpreted results of experiments; ZMH, DAS, JZ prepared figures; ZMH, JZ, DAS drafted manuscript; ZMH, JLL, LD, PKS, JZ, DAS edited, reviewed, and approved final version of manuscript.

## Notes

### Competing Interest Statement

The authors have declared no competing interest.

### Clinical Trial

NCT03078088

### Author Declarations

The University of Iowa IRB as well as the FDA approved the studies.

