## Supplemental Data for "Effects of THAM Nasal Alkalinization on Airway Microbial Communities: A Pilot Study"

### Suppl. Fig. 1

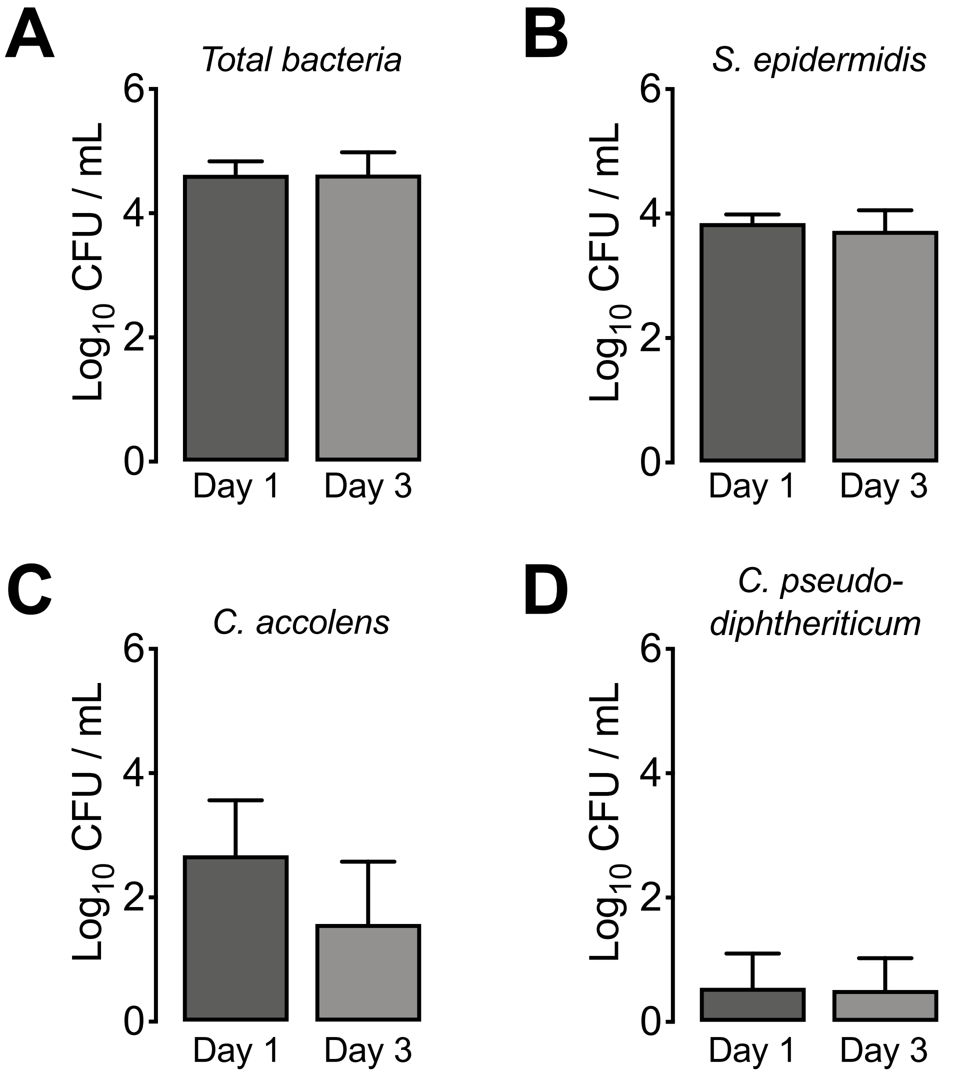

**Supplemental Figure 1. Human Nasal Bacterial Abundance and Composition are Stable.** Quantification of (**A**) total nasal bacteria, (**B**) *S. epidermidis*, (**C**) *C. accolens*, and (**D**) *C. pseudodiphtheriticum* in human subjects. Both nares were swabbed and bacterial counts quantified using standard microbiology techniques on day 1 and day 3. n=6 subjects. Bars are mean ± SEM

Suppl. Fig. 2

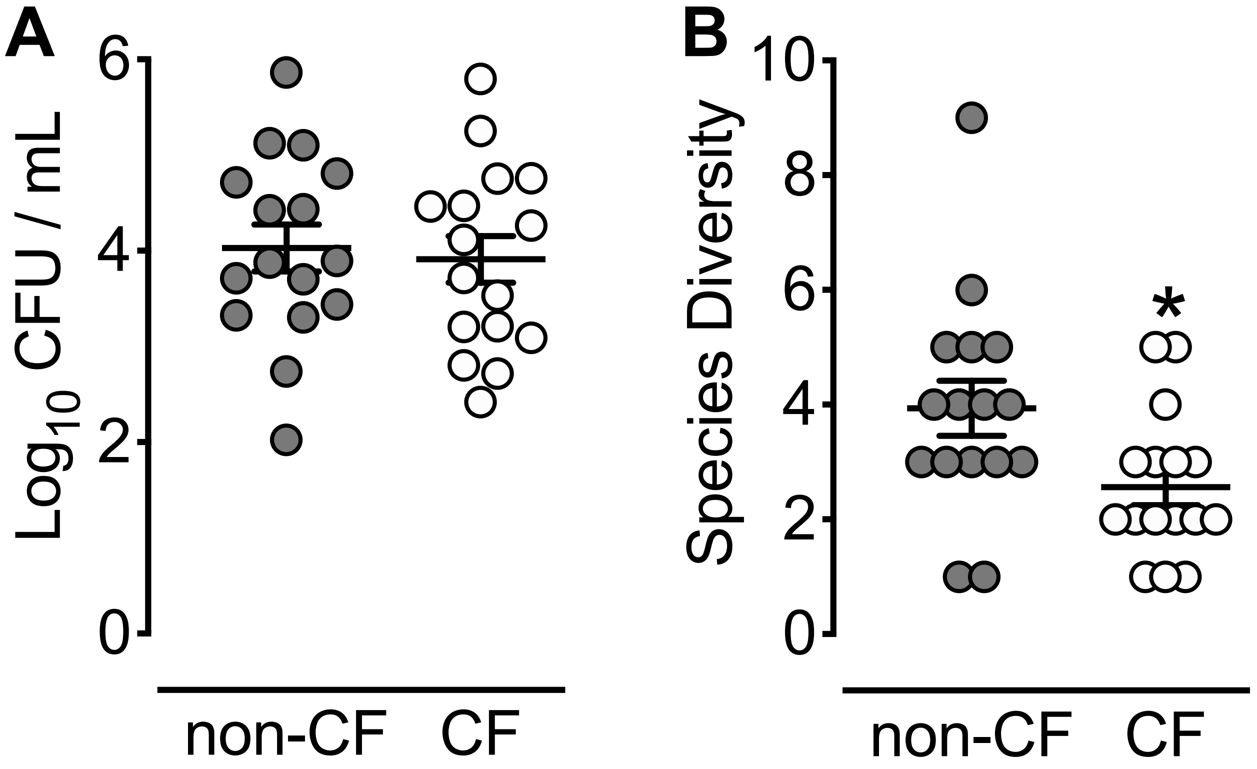

**Supplemental Figure 2. Baseline Nasal Swab Bacterial Counts and Species Diversity in Non-CF and CF Subjects.** (**A**) Bacterial counts obtained with nasal swabs. (**B**) Bacterial species diversity. Diveristy was determined by summing individual bacterial species from each subject. Each data point indicates nasal swab data from a different subject. Horizontal bars represent mean ± SEM. * denotes $P < 0.05$.

Suppl. Table 1

| Participant Compliance and Adverse Events | | | | |
|---|---|---|---|---|
| | non-CF | | CF | |
| **Participant Compliance** | | | | |
| Saline, THAM (SD) | 23.3 (1.5), 23.4 (1.6) | | 23.9 (0.25), 23.9 (0.25) | |
| total # doses = 24 | | | | |
| | non-CF | | CF | |
| **Adverse Events** | Saline | THAM | Saline | THAM |
| total number (%) | 1 (6.25%) | 1 (6.25%) | 3 (18.75%) | 1 (6.25%) |
| **Mild** | 1 | 1 | 3 | 0 |
| Cough | 0 | 0 | 1 | 0 |
| Chest tightness | 1 | 0 | 0 | 0 |
| Shortness of breath | 0 | 0 | 2 | 0 |
| Stuffy nose | 0 | 0 | 0 | 0 |
| Sneezing | 0 | 1 | 0 | 0 |
| **Moderate** | 0 | 0 | 0 | 1 |
| Back pain | 0 | 0 | 0 | 1 |
| **Serious** | 0 | 0 | 0 | 0 |

### Suppl. Table 2

| Most Frequently Isolated Bacterial Species | |
|---|---|
| **non-CF: *Gram (+)* bacteria** | **CF: *Gram (+)* bacteria** |
| *Corynebacterium accolens* | *Corynebacterium accolens* |
| *Corynebacterium pseudodiphtheriticum* | *Corynebacterium tuberculostearicum* |
| *Corynebacterium tuberculostearicum* | *Enterococcus faecalis* |
| *Staphylococcus aureus* | *Rhodotorula mucilaginosa* |
| *Staphylococcus capitis* | *Staphylococcus aureus* |
| *Staphylococcus epidermidis* | *Staphylococcus capitis* |
| *Staphylococcus haemolyticus* | *Staphylococcus epidermidis* |
| *Staphylococcus hominis* | *Streptococcus oralis* |
| *Staphylococcus pasteuri* | *Streptococcus parasanguinis* |
| *Staphylococcus warneri* | *Streptococcus sanguinis* |
| **non-CF: *Gram (–)* bacteria** | **CF: *Gram (–)* bacteria** |
| *Actinomyces oris* | *Achromobacter xylosoxidans* |
| *Escherichia coli* | *Klebsiella pneumonia* |
| *Haemophilus parainfluenzae* | *Pantoea eucrina* |
| *Klebsiella pneumonia* | *Pseudomonas aeruginosa* |
| *Moraxella nonliquifaciens* | *Serratia marcescens* |
